# Description and clinical validation of a real-time AI diagnostic companion for fetal ultrasound examination

**DOI:** 10.1101/2021.05.25.21257630

**Authors:** Julien J. Stirnemann, Remi Besson, Emmanuel Spaggiari, Sandra Rojo, Frederic Loge, Helene Peyro-Saint-Paul, Stephanie Allassonniere, Erwan Le Pennec, Yves Ville

**Affiliations:** Obstetrics and maternal-fetal medicine, Necker - Enfants Malades Hospital, AP-HP and EA7328 université de Paris, IMAGINE Institute; School of Medicine, université de Paris, INRIA EPI HEKA, INSERM UMR1138, Sorbonne université; Center for Applied Mathematics, Ecole Polytechnique. Institut Polytechnique de Paris; Department of Histology-Embryology and Cytogenetics, Unit of Embryo & Fetal Pathology, Necker-Enfants Malades Hospital, AP-HP and Paris Descartes University; INSERM UMR 1163, IMAGINE Institute; SONIO SAS, 24 rue du Faubourg Saint-Jacques, 75014 Paris; Xpop, INRIA Saclay

**Author notes:** **Correspondence:** Pr Julien STIRNEMANN, Maternité, Hôpital Necker Enfants Malades, 149 rue de Sevres 75015 Paris. **Funding:** None. **IRB :** CERAPHP.5 #00011928.

## Abstract

**Objective:** To describe a real-time decision support system (DSS), named SONIO, to assist ultrasound-based prenatal diagnosis and to assess its performance using a clinical database of precisely phenotyped postmortem examinations.

**Population and Methods:** This DSS is knowledge-based and comprises a dedicated thesaurus of 294 syndromes and diseases. It operates by suggesting, at each step of the ultrasound examination, the best next symptom to check for in order to optimize the diagnostic pathway to the smallest number of possible diagnoses. This assistant was tested on a single-center database of 251 cases of postmortem phenotypes with a definite diagnosis. Adjudication of discordant diagnoses was made by a panel of external experts. The primary outcome was a target concordance rate >90% between the postmortem diagnosis and the top-7 diagnoses given by SONIO when providing the full phenotype as input. Secondary outcomes included concordance for the top-5 and top-3 diagnoses; We also assessed a “1-by-1” model, providing only the anomalies sequentially prompted by the system, mimicking the use of the software in a real-life clinical setting.

**Results:** The validation database covered 96 of the 294 (32.65%) syndromes and 79% of their overall prevalence in the SONIO thesaurus. The adjudicators discarded 42/251 cases as they were not amenable to ultrasound based diagnosis. SONIO failed to make the diagnosis on 7/209 cases. On average, each case displayed 6 anomalies, 3 of which were considered atypical for the condition. Using the ‘full-phenotype’ model, the success rate of the top-7 output of Sonio was 96.7% (202/209). This was 91.9% and 87.1% for the top-5 and top-3 outputs respectively. Using the “1-by-1” model, the correct diagnosis was within the top-7, top-5 and top-3 of SONIO’s output in 72.4%, 69.3% and 63.1%.

**Conclusion:** Sonio is a robust DSS with a success-rate >95% for top-7 ranking diagnoses when the full phenotype is provided, using a large database of noisy real data. The success rate over 70% using the ‘1-by-1’ model was understandably lower, given that SONIO’s sequential queries may not systematically cover the full phenotype.

## INTRODUCTION

The discovery of a fetal malformation or anomaly during prenatal ultrasound examination occurs in around 2-3% of all pregnancies (1). Irrespective of its own severity, the diagnosis of one anomaly raises the question of whether this is an isolated finding or else it is associated with other anomalies within chromosomal or genetic syndromes that weigh negatively on its prognosis. There are over 9,000 rare diseases in the ORPHANET database (2), of which a few hundred express a consistent phenotype identifiable on prenatal ultrasound. However, the diversity of those syndromes and of many of their constitutive, often non-specific, features escape the knowledge of most specialists in prenatal diagnosis. Concurrently, ultrasound technology and technicity, driven by a growing demand, has reached a high level of details, raising the expectations of pregnant women for the assessment of the fetal development. There is therefore a discrepancy between the intractable complexity of rare diseases and the universal availability of powerful prenatal imaging technologies such as ultrasound. The medical uncertainty generated by this discrepancy, fueled by growing legal pressure, leads to an increasing number of ultrasound examinations per woman, without improving the diagnostic performance (3). This medical uncertainty is likely to affect the cost of prenatal screening for fetal anomalies and to generate unnecessary parental anxiety. The impact of the suspicion of a fetal anomaly is known to undermine the attachment of the pregnant woman to her pregnancy with a significant and prolonged impact even when reaching a favorable prognosis (4).

Given the growing complexity of medical knowledge and the increasing availability of data sources, decision support systems (DSS) have been developed to assist clinicians in their decision-making, particularly for rare diseases diagnosis (5,6). However, to our knowledge, such systems have not been developed for the specific needs of fetal ultrasound. Online search engines that link given fetal ultrasound phenotypes to specific diseases do exist (7), but such solutions implicitly assume that all possible fetal symptoms have been checked for and that the phenotype is therefore “complete” - which is usually not the case. Furthermore, such systems do not offer real-time assistance during ultrasound examination. Similar engines exist for postnatal clinical phenotypes, such as ‘Phenomizer’ developed with HPO and Orphanet (8).

We present the performance of a real-time DSS, named SONIO (9), that operates by suggesting, at each step of the ultrasound examination, the best next symptom to check for, in order to optimize the diagnostic pathway to the smallest number of possible diagnoses. This DSS is knowledge-based (10) and comprises a dedicated thesaurus constructed from several sources and two dedicated algorithms. This assistant was tested on a single-center database of postmortem phenotypes for which the rare disease diagnosis is certain.

## METHODS

### 1. Description of the Sonio DSS

#### Thesaurus of anomalies and syndromes

The sources of the Sonio thesaurus are Orphanet (2) and CRAT (11). Orphanet is an open source database of over 9,000 rare congenital disorders, produced by a consortium of 40 countries. In Orphanet, rare diseases are defined according to the European Union Regulation on Orphan Medicinal Products criteria, i.e. a disease that affects less than 1/2000 persons in the European population. A disorder in the database can be a disease, a syndrome, a morphological or a biological anomaly or a clinical condition. Each disorder or disease can be accessed using its name, ORPHAcode, gene symbol/name, OMIM number and classification (Online Mendelian Inheritance in Man), or ICD-10 (10th edition of WHO’s International Classification of Disease). The CRAT (‘Centre de référence des agents tératogènes’) is the French teratogenicity reference centre covering drugs, chemicals, radiations, and pathogens. It is maintained and updated by the Department of Pharmacology at Trousseau Hospital in Paris.

These 2 databases were screened according to the following criteria:

- conditions with a prevalence > 1 in a million births
- with a prenatal onset
- with visible anomalies on fetal ultrasound

The results were reviewed by a panel of experts from the Fetal medicine unit at Necker Hospital in Paris. This selection yielded 294 conditions extracted from the >9000 diseases listed in ORPHANET.

The SONIO’s thesaurus of phenotypic anomalies was built using the Human Phenotype Ontology (HPO) that provides a standardized vocabulary of phenotypic abnormalities encountered in human disease (12,13). Each term in the HPO describes a phenotypic abnormality, such as “Atrial septal defect”. The description of prenatal ultrasound features comprises all commonly accepted synonym terms used to name and characterize each anomaly.

Each disorder was also related to its possible identification by either standard prenatal karyotype or by prenatal comparative genomic hybridization (array-CGH)

The final Sonio’s thesaurus comprised 294 syndromes and disorders (Table 4) with a prevalence of between 1/400 (Down syndrome) and very rare (i.e. for which the prevalence is unknown but for which only case reports exist). Each of these disorders was linked to a total 668 possible ultrasound anomalies.

#### Description of the software algorithm

The workflow of the SONIO DSS is to start from a call anomaly or a risk factor, and displays at each iteration:

⍰ a number of visible abnormalities in the anatomical area being explored that could quickly guide the diagnosis
⍰ the next anatomical areas to verify for any associated anomalies
⍰ contextual questions (if necessary) about possible risk factors
⍰ the serial probability of the different diagnoses

The SONIO main software is built on a combination of two algorithms: one for the diagnosis of a syndrome given the phenotype and the second to suggest the next ultrasonographic anomaly to look for (14).

The diagnosis algorithm consists of performing a Bayes formula commonly used for building probabilistic reasoning systems (15) particularly in medical applications (16) with several algorithmic refinements. It is fitted to work with medical ontologies (12,17), since the anomalies are related to each other by the tree structure of HPO. This means that some anomalies are “ascendants” of others, i.e. they describe a more general concept. In such a case, it is necessary to direct the information through the ontology graph to estimate more precisely the probabilities of the anomalies given the corresponding syndrome. The algorithm is also able to process contextual information, such as fetal gender, gestational age, etc. that change the probability of the syndromes or of the anomalies. Then, the algorithm can deal with the causal links between the anomalies. Indeed, some anomalies, for example Pierre-Robin sequence, are more complex entities which include other anomalies such as cleft palate or micro retrognatia. For such cases we need an AND gate logic with rule-based methods that is not encapsulated in the HPO ontology and which is one of the legacies of the expert systems of the 1980s (18). The second main algorithm directly uses this diagnosis algorithm to suggest the next anomalies to look for. This algorithm mainly consists of a decision tree algorithm based on average information gain (19,20). Nevertheless, several refinements have been necessary to produce acceptable diagnostic pathways for the users. First, we accounted for the ergonomics of the ultrasound examination with, for example, no back and forth between anatomical zones. The software’s line of reasoning must also appear logical to the user in order to be adopted. This is why the software is designed to first perform causal reasoning, i.e starts from the input sign and looks for a mechanical cause, before starting to reason by association of malformations to diagnose a syndrome.

### 2. Design and setting of the validation study

This was a prospectively designed study performed on a cohort of 251 cases extracted from the Necker database of fetal post mortem pathology, with at least one sonographic anomaly and a definite diagnosis. A formal protocol was established for each of the key steps of the study prior to its implementation:

- Curation and annotation of the postmortem phenotypes and diagnoses that define the validation database
- Running SONIO on the postmortem phenotypes of the validation database
- Definition of success criteria
- Adjudication using a panel of external experts for cases on which Sonio and the post mortem diagnosis were discordant: was this a true failure of Sonio DSS or was the diagnosis altogether impossible because of a truly atypical phenotype based upon the aforementioned Orphanet (i.e. the postmortem diagnosis was fortuitously made only by using wide genetic testing)?

#### Curation and annotation of the validation database of postmortem phenotypes

The validation database was extracted from the Necker database of fetal pathology using the following criteria: the syndrome is present in the SONIO thesaurus; from the pathology report, ultrasound detectable abnormalities can be identified (i.e. symptoms found in the SONIO thesaurus); the diagnosis is certain, confirmed by genetic testing or clinically by a geneticist and identified by an ORPHA ID (Figure 1). Additional data was collected, including gestational age at the time of diagnosis and a set of known risk factors (e.g. gender, recent Zika Infection etc.)

This search yielded 251 cases (comprising 1689 ultrasound anomalies). The validation database covered 96 syndromes of the 294 (32.65%) contained in the Sonio thesaurus. In terms of prevalence these 96 syndromes represent 79% of the 294 syndromes of the Sonio thesaurus. The postmortem diagnoses cover genetic single-gene, chromosomal, infectious and toxic/teratogenic etiologies.

This study was approved by the local ethics committee of the Necker hospital (IRB registration : CERPAHP.5 #00011928).

#### Emulating tools to run SONIO on postmortem phenotypes

To mimic the use of Sonio in real-life clinical ultrasound, the software was applied to the postmortem phenotypes using two emulating models. We call ‘emulation’, a scenario of application. The 2 models were defined as follows :

⍰ “full phenotype”: in this model, for each case, all risk factors and observed anomalies are provided to SONIO as a single input. This model provides an estimate of the crude performance of the system. Several scenarios were emulated in each case using different assumptions for gestational age (when not filled in the case), or previous CGH array testing whenever this data was missing.
⍰ “1-by-1”: in this model, the system is primed with a randomly selected anomaly as an input and sequentially questions the presence or absence of specific anomalies, until all the anatomical regions of the fetus have been checked. A region is declared valid as soon as all the anomalies recommended by SONIO on this particular region have been checked for as present or absent respectively. However, the 1^st^ anomaly could not be an atypical sign, i.e. a sign with <5% probability of association with the disease.

#### Definition of success criteria

The primary outcome was based upon the use of the “full-phenotype” emulation, with a target concordance rate > 90% between the postmortem diagnosis and the top-7 diagnoses given by SONIO (i.e. the postmortem diagnosis was within the 7 most likely diagnoses suggested by SONIO). Secondary outcomes included concordance for the top-5 and top-3 diagnoses; concordance using the “1-by-1” model; number of steps to reach the final list of diagnoses; number of discordant cases.

#### Management of discordances between Sonio output and postmortem diagnosis

An adjudication protocol was designed for discordances between SONIO and postmortem diagnosis. Each case was adjudicated by 2 of 3 independent external fetal medicine experts (FA, AG, TS). A third expert was asked to referee disagreements between two experts. For each case to be adjudicated, each one of the experts was given the complete phenotype and was asked for the most likely diagnosis, blinded to the postmortem diagnosis as well as to the Sonio output. The expert diagnosis was then compared to the SONIO and postmortem diagnosis with the following 4 possible decisions : i. SONIO and postmortem agree, which happened when the initial discordance was based on synonym terms or two subtypes of the same disease; ii. The diagnosis was achievable given the phenotype but SONIO failed; iii. the diagnosis was impossible given the ultrasound phenotype (unspecific/atypical phenotype); iv. The postmortem diagnosis was incorrect.

## RESULTS

The thesaurus of the DSS comprises 294 syndromes and diseases with a prevalence of 1/400 to 1 in a million and 668 prenatal ultrasound anomalies. The validation database of 251 postmortem phenotypes, ranging from 12 weeks to 40 weeks, covered 96 syndromes or diseases (73 single-gene, 17 chromosomal, 2 infectious, 4 toxic/teratogenic). Each case displayed 6 anomalies on average (min=2, max=21), 3 of which being considered atypical of the disease (Table 1). The validation database covered 32.65% (96/294) of the syndromes of SONIO’s thesaurus but 79% of their overall cumulative prevalence. The array-CGH, karyotype, FISH or gene-sequencing was abnormal in only 107/251 (42%) of the cases. The other postmortem diagnoses were based solely on clinical assessment.

**Table 1.** Baseline description of the validation dataset and SONIO emulations

|  |  | Total | Median | Min | Max |
| --- | --- | --- | --- | --- | --- |
| Nb of scenarios emulated for each case | full phenotype | 464 | 2 | 1 | 4 |
|  | 1-by-1 | 1210 | 4 | 0 | 20 |
| Prior adjudication | Number of anomalies per case |  | 6 | 1 | 20 |
|  | Number of atypical anomalies per case |  | 3 | 0 | 19 |
| Post adjudication | Number of anomalies per case |  | 6 | 1 | 20 |
|  | Number of atypical anomalies per case |  | 2 | 0 | 19 |
| Number of steps to diagnosis starting from one anomaly |  |  | 9 | 4 | 24 |

A total of 464 scenarios were emulated on SONIO with the ‘full phenotype’ model. With the “1-by-1” model 1,210 emulations were run, requiring 9 steps on average (i.e. 9 questions asked by SONIO) before reaching a final list of potential diagnoses. Concordance between the top-7 SONIO diagnoses and postmortem was achieved in 199 cases for the full-phenotype mode, and 52 cases required adjudication by the external panel of experts (Figure 2). The adjudication excluded 42 diagnoses that were considered to be unachievable with the available information, leaving 209 cases for which the diagnosis is achievable using the postmortem phenotype alone. Three cases were considered concordant after adjudication, essentially because of synonym names. In 7/209 SONIO failed to achieve the diagnosis within the top-7. Using the ‘full-phenotype’ model, the success rate of the top-7 output of SONIO was 96.7% (202/209) with a confidence interval of [93.8; 99.6]. This was 91.9% and 87.1% for the top-5 and top-3 outputs respectively (Table 2). Using the “1-by-1” model without the CGH array results, the correct diagnosis was within the top-7, top-5 and top-3 of SONIO’s output in 72.4%, 69.3% and 63.1% of the case respectively. Adding the information of a normal CGH array in non-chromosomal syndromes yielded success rates of 74%, 71% and 65% within the top-7, top-5 and top-3 respectively (Table 3).

**Table 2.** Concordance table for validation cases with the ‘full phenotype’ model.

| Rank of true syndrome in suspects | Pre ADJUDICATION |  | POST ADJUDICATION |  |
| --- | --- | --- | --- | --- |
|  | N / N cases | Proportion | N / N cases | Proportion |
| In top-7 | 198 / 251 | 79% | 202 / 209 | 97% |
| In top-5 | 191 / 251 | 76% | 192 / 209 | 92% |
| In top-3 | 181 / 251 | 72% | 182 / 209 | 87% |

**Table 3.** Concordance table for validation cases with the ‘1-by-1’ model.

| CGH OPTION | Rank of true syndrome in suspects | PreADJUDICATION |  | POST ADJUDICATION |  |
| --- | --- | --- | --- | --- | --- |
|  |  | N/N cases | % | N/N cases | % |
| CGH was normal | In top-7 | 406 / 563 | 72% | 397 / 534 | 74% |
|  | In top-5 | 385 / 563 | 68% | 378 / 534 | 71% |
|  | In top-3 | 353 / 563 | 63% | 349 / 534 | 65% |
| CGH not done | In top-7 | 446 / 647 | 69% | 441 / 609 | 72% |
|  | In top-5 | 427 / 647 | 66% | 422 / 609 | 69% |
|  | In top-3 | 388 / 647 | 60% | 384 / 609 | 63% |

The details of the 7 cases considered as a SONIO failure by adjudication can be found in Table 4. The details of the 42 cases that were excluded by the adjudicators can be found in Supplementary Table S1, highlighting rare chromosomal anomalies detectable by CGH or mosaics and the importance of careful abnormality description and coding while analyzing clinical cases. More details and explanations on these cases can be found in supplementary material.

**Table 4.**
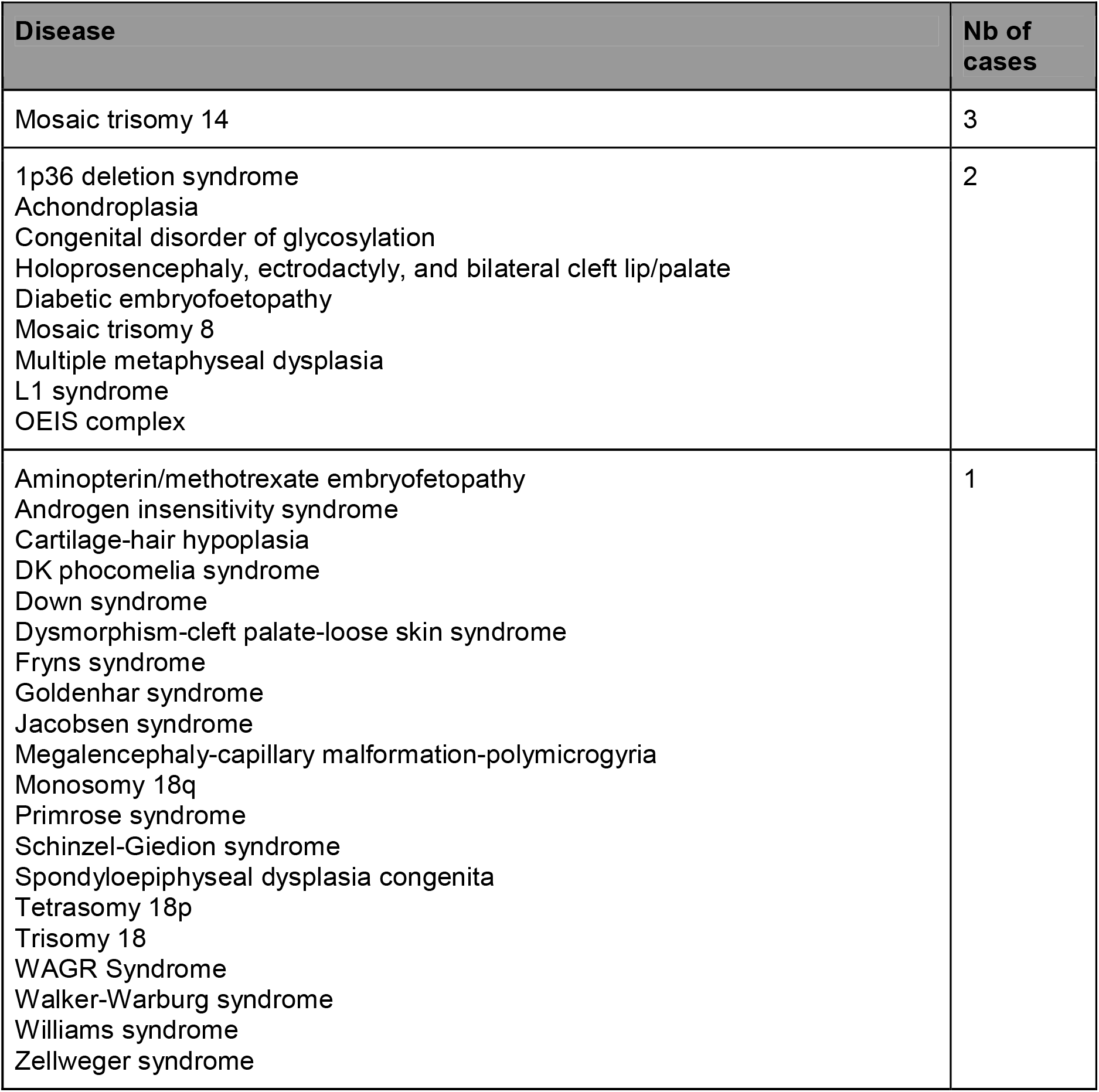
List of the diseases affecting the 42 cases labelled by adjudicators as “Unlikely to be diagnosed based on available phenotype”

| Disease | Nb of cases |
| --- | --- |
| Mosaic trisomy 14 | 3 |
| 1p36 deletion syndrome<br>Achondroplasia<br>Congenital disorder of glycosylation<br>Holoprosencephaly, ectrodactyly, and bilateral cleft lip/palate<br>Diabetic embryofetopathy<br>Mosaic trisomy 8<br>Multiple metaphyseal dysplasia<br>L1 syndrome<br>OEIS complex | 2 |
| Aminopterin/methotrexate embryofetopathy<br>Androgen insensitivity syndrome<br>Cartilage-hair hypoplasia<br>DK phocomelia syndrome<br>Down syndrome<br>Dysmorphism-cleft palate-loose skin syndrome<br>Fryns syndrome<br>Goldenhar syndrome<br>Jacobsen syndrome<br>Megalencephaly-capillary malformation-polymicrogyria<br>Monosomy 18q<br>Primrose syndrome<br>Schinzel-Giedion syndrome<br>Spondyloepiphyseal dysplasia congenita<br>Tetrasomy 18p<br>Trisomy 18<br>WAGR Syndrome<br>Walker-Warburg syndrome<br>Williams syndrome<br>Zellweger syndrome | 1 |

Figures 3, 4 and 5 provide visual representations of these results. Concerning the figure 3, the y-axis gives the name of the anomalies that were used as an entry point in the emulation of the software, on the x-axis are the names of the syndromes to be found as diagnostic. One square can refer to several scenarios emulated on Sonio for the same syndrome and entry point, since there may be several clinical cases with the same diagnosis. For that reason we represent the average results, the closer this square is to yellow, the closer the software is to having systematically found the right diagnosis starting from the considered entry point. The lowest line shows the results of the full-phenotype (all the file) option. Thus this figure shows the correspondence between the cases failing on all the file (in the right end of the axis) with the cases failing on the “1-by-1” mode (upper right frame of the graph).

Figure 4 represents another visualization of the performance of the “all the file” on the y-axis and the performance of the “1-by-1” mode on the x-axis. The group of syndromes in the upper right-hand frame of the graph are systematically diagnosed by the software whether one gives the whole phenotype at once or follows the decision tree. The cases at the lower part of the graph are those that went to adjudication. Finally, the cases at the upper left-hand box of the graph were correctly diagnosed by Sonio when the whole phenotype is provided but are not when the decision tree is followed. The intervariability of the performance *per* diagnostic can be analyzed in more detail at Figure 5

## DISCUSSION

### Main results

In this study, we present the description and clinical validation of a diagnostic support system for fetal ultrasound on real data, using a database of postmortem phenotypes with a certain diagnosis. This study showed that SONIO identified the post-mortem diagnosis within the 7 most probable diagnoses in >95% of cases when the full phenotype is provided. These results, together with the >91% concordance rate for top-5 list, show that SONIO was remarkably robust to noise, given that, on average, half of the anomalies were considered atypical for the diagnosis. In addition, although it did slightly improve the results, providing information that array-CGH was normal in non-chromosomal syndromes was of little help. The performance of SONIO was expectedly lower when using the ‘1-by-1’ model, although successful in over 70% of the cases: indeed, under this model, the complete phenotype may not have been provided to SONIO in the end, since only the symptoms that SONIO has asked to be checked are accounted in the diagnostic pathway. This model is closer to what one can expect in real-life than the ‘full phenotype’ model.

### Interpretation

Faviez et al. recently reviewed the performance of diagnostic DSSs for rare diseases (6). The systems presented in this review rely on post-hoc postnatal phenotyping and do not provide real-time assistance. In the 9 systems for which performance was reported, the success-rate of the top-10 diagnoses were uniformly spread from 32% to 99% (22). SONIO, which incorporates the additional complexity of real-time processing, therefore appears as an effective tool with a success rate in the highest performance range of the available rare disease DSS tools, while using the top-7 diagnoses, a more stringent criterion than the top-10 used by others. This is particularly relevant, given that reported performances rely rarely on real data for validation, but generally use simulated / in silico validation.

### Strengths and limitations

The strengths of this study are:

- A unique database: the database contains 251 cases (96 diseases). However, this is likely to approximate 79% of the combined prevalence of diseases encountered by sonographers during prenatal foetal US examination by summing-up individual prevalences reported by ORPHANET. This makes it therefore suitable for use in clinical practice. Furthermore, compared to real-life prenatal diagnosis, this database is enriched with the most challenging rare, non-chromosomal diagnoses since chromosomal defects, generally confirmed prenatally with array-CGH generally do not require a postmortem examination
- The emulation models describe two different aspects of the performance of SONIO: the ‘full phenotype’ model describes the performance of the database and diagnostic search engine, while the ‘1-by-1’ emulates the real-life use
- The external independent adjudication of discordant cases

The limitations of this study are:

- The validation dataset is based on the postmortem data of a single center: despite the high coverage of possible diagnoses, it is incomplete. It may also be biased; through preferential selection of a subset of conditions for post-mortem examination; it may also originate from a center-specific postmortem phenotype assessment.
- When using SONIO during real-life fetal ultrasound examination, the software sequentially suggests anomalies to look for, but the interpretation of the presence or absence of an anomaly on the ultrasound image is made by the clinician. In our validation setting, we provided an accurate phenotype established by the postmortem examination, which therefore did not incorporate potential human error in the interpretation of ultrasound imaging and risk factors (23).
- In the “1-by-1” model, the first input is always a characteristic symptom of the disease (i.e. that is present in at least 5% of patients affected with the syndrome). We therefore did not assess the software when the first symptom is an atypical sign.

### Clinical impact of this study

The knowledge by one individual of the precise prenatal phenotype of over 300 congenital disorders or syndromes as well the respective prevalence and specificity of each one of their constitutive anomalies is unlikely. On average, around 50% of those undergo a complete diagnosis before birth. We therefore believe that the results of this validation process strongly support that SONIO’s algorithm is a significant augmentation to the diagnostic capability of ultrasound-based prenatal diagnosis.

Artificial intelligence in this field of fetal medicine is developing in two main directions, the recognition of standardized ultrasound images for assessment of normality or biometrics (24) and that of diagnostic DSSs (6). SONIO belongs to the latter. The clinical strengths of SONIO lie in: i) the quality of its constitutive thesaurus which is a prenatal adaptation of Orphanet, the most recognized and up-to-date registry of rare congenital disorders; ii) the incorporation of context information and risk factors (e.g. previous genetic testing, gestational age, sex etc.) in the algorithms; iii) Its ability to work in real-time at the patient’ side in the presence of a call anomaly or a risk factor by prompting the practitioner to look for anomalies that would possibly have been overlooked otherwise; iv) maintaining the diagnostic process within acceptable duration and number of steps.

This study adds convincing external validation results on real data to this list. We acknowledge such a validation, based on postmortem phenotypes, while providing the best possible reference diagnosis quality, does not provide the same information as a prospective controlled, possibly randomized, validation trial. However, such a trial is unlikely to ever be implemented as the prevalence of diseases covered by SONIO would require either an unrealistic number of pregnancies to examine or an unrealistic study length. This limitation, as well as the possible false-positive diagnoses that SONIO may generate in the absence of congenital disease, will be addressed in further studies.

## Conclusion

In conclusion, SONIO has developed an effective real-time diagnostic DSS for specialists involved in prenatal diagnosis that is likely to improve the quality of their work as well as provide continuous medical education. We believe that the confidence gained by the physician could also improve the patient-physician interaction.

## Data Availability

The validation data will be made partly available, upon request.
The algorithms and thesaurus of the SONIO DSS are the property of the SONIO company

## ACKNOWLEDGMENTS

We thank François Audibert, Annegret Geipel, Thomas Schramm for their implication in the adjudication process.

## LEGENDS TO TABLES AND FIGURES

Figure 1: Example of an annotated postmortem phenotype

Figure 2. Flow chart of the study population

Figure 3 - Percentage of scenarios emulated in sonio (blue - green - yellow scale, yellow being the best) indicating the correct diagnosis within top-7 suggestions, by Syndrome (x axis) and anomaly of entry (y axis). In y axis were added two categories, identified on the bottom left of the graph: starting with the whole file (“All the file”) and starting from a specific anomaly (“One anomaly”). E.g. the Holt-Oram syndrome cases were not found giving the whole life whilst always found starting from single anomalies. Note that syndromes were sorted first by performance on the whole file and second on performance starting from a single anomaly, hence syndromes close to the left are the ones which were most often correctly found. Note also that anomalies (aside the grouping categories) were sorted by their average performance as well. Gray cells have no data support i.e. no simulation for the given syndrome and anomaly of entry at their coordinates.

Figure 4 - Percentage of scenarios emulated in sonio indicating the correct diagnosis within the top-7 starting from one anomaly (in x axis) versus giving the whole file (in y axis) by syndrome (regrouping several cases). BOR and Joubert Syndromes are in coordinate (1,1), indicating that we managed to find those syndromes in our top-7 suggestions whether starting with a single anomaly or the whole file. The majority of our cases perform very well when using the whole life as we have seen previously (see the concentration of points around y = 1). A few files have overall poor performance.

Figure 5 - Percentage of scenarios emulated in sonio indicating the correct diagnosis within the top-7 starting from one anomaly (right boxplot) versus giving the whole file (left boxplot) by syndrome (each facet). Achondroplasia cases perform well when all the file is given, whilst starting with an anomaly gives a very volatile performance i.e. depending on the anomaly of entry, performance ranges 10% to 60% chances of finding the right diagnoses in top-7. This graph is an alternative and slightly more detailed view of the preceding figure.

## REFERENCES

1. EUROCAT [Internet]. [cited 2010 Sep 28]. Available from: http://www.eurocat-network.eu/

2. Orphanet, The portal for rare diseases and orphan drugs [Internet]. [cited 2021 May 18]. Available from: http://www.orpha.net/consor/www/cgi-bin/index.php?lng=EN

3. Ferrier C, Dhombres F, Khoshnood B, Randrianaivo H, Perthus I, Guilbaut L, et al. Trends in resource use and effectiveness of ultrasound detection of fetal structural anomalies in France: a multiple registry-based study. BMJ Open. 2019 Feb;9(2):e025482.

4. Api O, Demir HN, Api M, Tamer I, Orbay E, Unal O. Anxiety scores before and after genetic sonogram. Arch Gynecol Obstet. 2009 Oct;280(4):553–8.

5. Gambhir S, Malik SK, Kumar Y. Role of Soft Computing Approaches in HealthCare Domain: A Mini Review. J Med Syst. 2016 Dec;40(12):287.

6. Faviez C, Chen X, Garcelon N, Neuraz A, Knebelmann B, Salomon R, et al. Diagnosis support systems for rare diseases: a scoping review. Orphanet J Rare Dis. 2020 Apr 16;15(1):94.

7. Porat S, de Rham M, Giamboni D, Van Mieghem T, Baud D. Phenotip - a web-based instrument to help diagnosing fetal syndromes antenatally. Orphanet J Rare Dis [Internet]. 2014 Dec 10 [cited 2021 May 14];9. Available from: https://www.ncbi.nlm.nih.gov/pmc/articles/PMC4268872/

8. The Phenomizer (Orphanet) - Clinical Diagnostics with Similarity Searches in Ontologies [Internet]. [cited 2021 May 20]. Available from: http://compbio.charite.de/phenomizer_orphanet/

9. Guided screening and diagnosis of congenital malformations - sonio sonio [Internet]. [cited 2021 May 20]. Available from: https://www.sonio.ai/

10. Montani S, Striani M. Artificial Intelligence in Clinical Decision Support: a Focused Literature Survey. Yearb Med Inform. 2019 Aug;28(1):120–7.

11. CRAT - Centre de référence sur les agents tératogènes chez la femme enceinte [Internet]. [cited 2021 May 19]. Available from: https://www.lecrat.fr/

12. Köhler S, Gargano M, Matentzoglu N, Carmody LC, Lewis-Smith D, Vasilevsky NA, et al. The Human Phenotype Ontology in 2021. Nucleic Acids Res. 2021 Jan 8;49(D1):D1207–17.

13. Human Phenotype Ontology [Internet]. [cited 2021 May 19]. Available from: https://hpo.jax.org/app/

14. Besson R. Decision making strategy for antenatal echographic screening of foetal abnormalities using statistical learning [Internet] [These de doctorat]. Université Paris-Saclay (ComUE); 2019 [cited 2021 May 19]. Available from: http://www.theses.fr/2019SACLX037

15. Probabilistic Reasoning in Intelligent Systems [Internet]. Elsevier; 1988 [cited 2021 May 19]. Available from: https://linkinghub.elsevier.com/retrieve/pii/C20090276094

16. Charniak E. The Bayesian Basis of Common Sense Medical Diagnosis. In: AAAI. 1983.

17. Dhombres F, Jouannic J-M, Jaulent M-C, Charlet J. Choix méthodologiques pour la construction d’une ontologie de domaine en médecine prénatale. 2010;13.

18. Jackson P. Introduction to expert systems [Internet]. undefined. 1986 [cited 2021 May 19]. Available from: https://www.semanticscholar.org/paper/Introduction-to-expert-systems-Jackson/719e4e1328be9487b33a13dc38b6120993999ed5

19. Quinlan JR. Induction of decision trees. Mach Learn. 1986 Mar 1;1(1):81–106.

20. Breiman L, Friedman J, Stone CJ, Olshen RA. Classification and Regression Trees. Taylor & Francis; 1984. 372 p.

21. Chen J, Xu H, Jegga A, Zhang K, White PS, Zhang G. Novel phenotype-disease matching tool for rare genetic diseases. Genet Med Off J Am Coll Med Genet. 2019 Feb;21(2):339–46.

22. Zemojtel T, Köhler S, Mackenroth L, Jäger M, Hecht J, Krawitz P, et al. Effective diagnosis of genetic disease by computational phenotype analysis of the disease-associated genome. Sci Transl Med. 2014 Sep 3;6(252):252ra123.

23. Smith-Bindman R, Hosmer WD, Caponigro M, Cunningham G. The variability in the interpretation of prenatal diagnostic ultrasound: Interpretation of prenatal diagnostic ultrasound. Ultrasound Obstet Gynecol. 2001 Apr;17(4):326–32.

24. Drukker L, Noble JA, Papageorghiou AT. Introduction to artificial intelligence in ultrasound imaging in obstetrics and gynecology. Ultrasound Obstet Gynecol Off J Int Soc Ultrasound Obstet Gynecol. 2020 Oct;56(4):498–505.

