## supplementary information for "Description and clinical validation of a real-time AI diagnostic companion for fetal ultrasound examination"

SUPPLEMENTARY MATERIAL

**Details regarding the discordant cases and adjudication process**

First, among the 42 cases labelled by adjudicators as “Unlikely to be found by Sonio as non-specific phenotype”, there are 14 cases with a chromosomic syndrome (33% of the total) These cases, in practice, have been incidentally diagnosed at karyotype motivated by a risk factor or a nonspecific malformation (Mosaic trisomy 14, Mosaic trisomy 8, 1p36, Tetrasomy 18p, …).

Another group of these 42 cases is formed of 26 genetic diseases (62% of the total), a part of them present a very unspecific phenotype, another part present a very atypical form of the syndrome and could be attributable to an encoding error when building the clinical database or to a rare form of the syndrome (L1 syndrome with an unilateral renal agenesis and a distal arthrogryposis, Achondroplasia without macrocephaly nor micromelia but with an arhinencephaly and a hypoplasia of the cerebellar vermis with agenesis of corpus callosum, etc...).

Among the 7 cases labelled as “True error of Sonio” by the adjudicators, one case of diabetic embryopathy was suggested to be Beckwith-Wiedemann syndrome. The maternal context had not been integrated beforehand in Sonio. A similar error was made with a case of fetal valproate syndrome although maternal treatment was known by Sonio, that presented with growth restriction and cardiac defect but also unexpected oligohydramnios and absence of other typical anomalies. A case of Holt-Oram was missed while presenting a typical abnormal thumb morphology and cardiac defect but failing to mention aplasia or hypoplasia of the radius and showing an unexpected preaxial hand polydactyly. A case of Simpson-Golabi-Behmel was typically large for gestational age with nephromegaly. Nevertheless, macrocephaly, macroglossia, polyhydramnios and hepatomegaly were missing and there were unexpected hypertelorism and cardiac defect (VSD with DORV and hypoplasia of the right ventricle). Finally a case of Walker-Warburg showed expected lissencephaly and abnormal orbital region but failed to show cerebellar vermis hypoplasia hydrocephalus or even ventriculomegaly. In addition, it showed definitely atypical anomalies in WW with micro-retrognathia, meningocele and a long philtrum.

Table S1. Diseases of the 7 cases labelled as “True error of Sonio” by adjudicators

| **Disease** | **Nb of cases** |
| --- | --- |
| Arthrogryposis multiplex congenita  Currarino syndrome  Diabetic embryofoetopathy  Fetal valproate syndrome  Holt-Oram syndrome  Simpson-Golabi-Behmel syndrome  Walker-Warburg syndrome | 1 |
